# Delineating the effect of sex hormone intake on immunity in cis and trans women with HIV

**DOI:** 10.1101/2023.05.09.23289654

**Authors:** Chloé Pasin, David Garcia Nuñez, Katharina Kusejko, Anna Hachfeld, Hélène Buvelot, Matthias Cavassini, Lauro Damonti, Christoph Fux, Begoña Martinez de Tejada, Julia Notter, Alexandra Trkola, Huldrych F. Günthard, Karoline Aebi-Popp, Roger D. Kouyos, Irene A. Abela, the Swiss HIV Cohort Study

## Abstract

**Background:** Although sex hormones are recognized to induce immune variations, little is known on the effect of exogenous sex hormone intake on immune responses in cis and trans women. Here, we aimed at quantifying how sex hormone intake affects HIV-1 immune markers in cis women (CW) and trans women (TW) with HIV.

**Methods:** We considered measurements of key HIV-1 immune markers (CD4, CD8, lymphocyte counts, and CD4:CD8 ratio) from cis men (CM), CW, and TW enrolled in the Swiss HIV Cohort Study. We modeled immune markers using linear mixed-effects models with an interaction between the variables “group” (CW, TW) and “with sex hormone intake” (yes/no). We conducted serum proteomics measurements of 92 inflammation markers on samples from 31 TW before and after sex hormone intake to assess the inflammation environment.

**Results:** We included 54’141 measurements from 3’092 CW and 83 TW sampled between 2015 and 2022, and 147’298 from 8’611 CM. Sex hormone intake was associated with significant distinct effects on CD4 and CD4:CD8 ratio between the different groups of women (p=0.0025 and 0.015). TW with sex hormone intake had significantly higher CD4 counts (median = 772 (1Q-3Q=520-1’006)) than without (median = 617 (1Q-3Q=426-892)). This increase was similar in magnitude to the difference in CD4 counts between CW and CM. None of the serum inflammation proteins showed significant concentration difference before and after sex hormone intake in TW.

**Conclusion:** This study highlights the need to consider the potential role of sex hormone intake in modulating the immune system among other biological and social factors, especially in TW in HIV.

## Introduction

Sex and gender differences in the immune response have been observed in many settings, and in general women mount more robust cellular and humoral immune responses to infection and vaccination compared to men (1-6). These differences are multifactorial and could be influenced by the immuno-modulatory effect of sex hormones (7-9), with oestrogens having a stimulating effect (10) while testosterone having a suppressive effect on immune function (11). Sex hormones have also shown a multilevel effect on chronic infections such as HIV-1 (12), where women have displayed lower viral loads (13-15) and higher CD4+ T cell counts (16, 17) than men following seroconversion, and higher levels of immune activation (18, 19). Women also tend to have a lower latent HIV-1 reservoir size (20-22) and oestrogens were found to inhibit HIV-1 transcription (23). Although sex hormones could play a role in HIV-1 acquisition risk (24), recent evidence suggest that hormonal contraception does not affect the risk of HIV-1 acquisition (25-28) or disease progression (29, 30).

However, the effect of sex hormones intake on HIV-1 immune markers dynamics has been insufficiently explored. Additionally, most of the findings previously mentioned were obtained by studying cis women (CW) versus cis men (“cis” referring to people whose gender identify matches the one assigned at birth), or without specifying this information. Trans women (TW, referring to people who were recorded male at birth but do not identify with being a man) are disproportionally affected by HIV-1 (31, 32) and constitute a distinct epidemiologic group compared to men having sex with men and cis women (32-34). TW are still under-represented in HIV-1 research; therefore, little is known on the immune effect of sex hormone intake in TW and more generally on the effect of hormonal therapy taken by TW on HIV-1 suppression and longer-term health outcomes (35-38). Additionally, concerns on drug-drug interactions between antiretroviral treatment (ART) and sex hormones might limit hormone prescription in women with HIV-1 (39, 40).

The aim of this study was to analyze sex hormone intake in CW and TW from the SHCS, and to delineate the effect of sex hormones on key HIV-1 markers in CW and TW with sex hormone intake or not. Furthermore, we aimed to characterize the potential immuno-modulatory effect of sex hormones in TW by measuring inflammatory protein levels before and after sex hormone intake using a high-throughput proteomics assay.

## Results

### Study population

Participants from the Swiss HIV Cohort Study (SHCS) attend biannual visits during which blood samples are drawn and the following laboratory measurements are realized: number of leucocytes, lymphocytes, CD3+, CD4+, and CD8+ T cells (cells/μL), HIV-1 viral load (copies/mL). We considered measurements realized after January 1^st^, 2015: from that date, the SHCS data contains systematic report of most comedications using the ATC codes nomenclature. We included in the analysis a total of 54’141 laboratory measurements between January 2015 and September 2022 from 954 CW younger than 40 years-old (CW<40), 2’365 CW between 40 and 60 years-old (CW 40-60), 573 CW older than 60 years-old (CW>60), and 83 TW (Figure 1). As a comparison we also considered similarly selected 147’298 laboratory measurements from 8’611 cis men.

**Figure 1:**
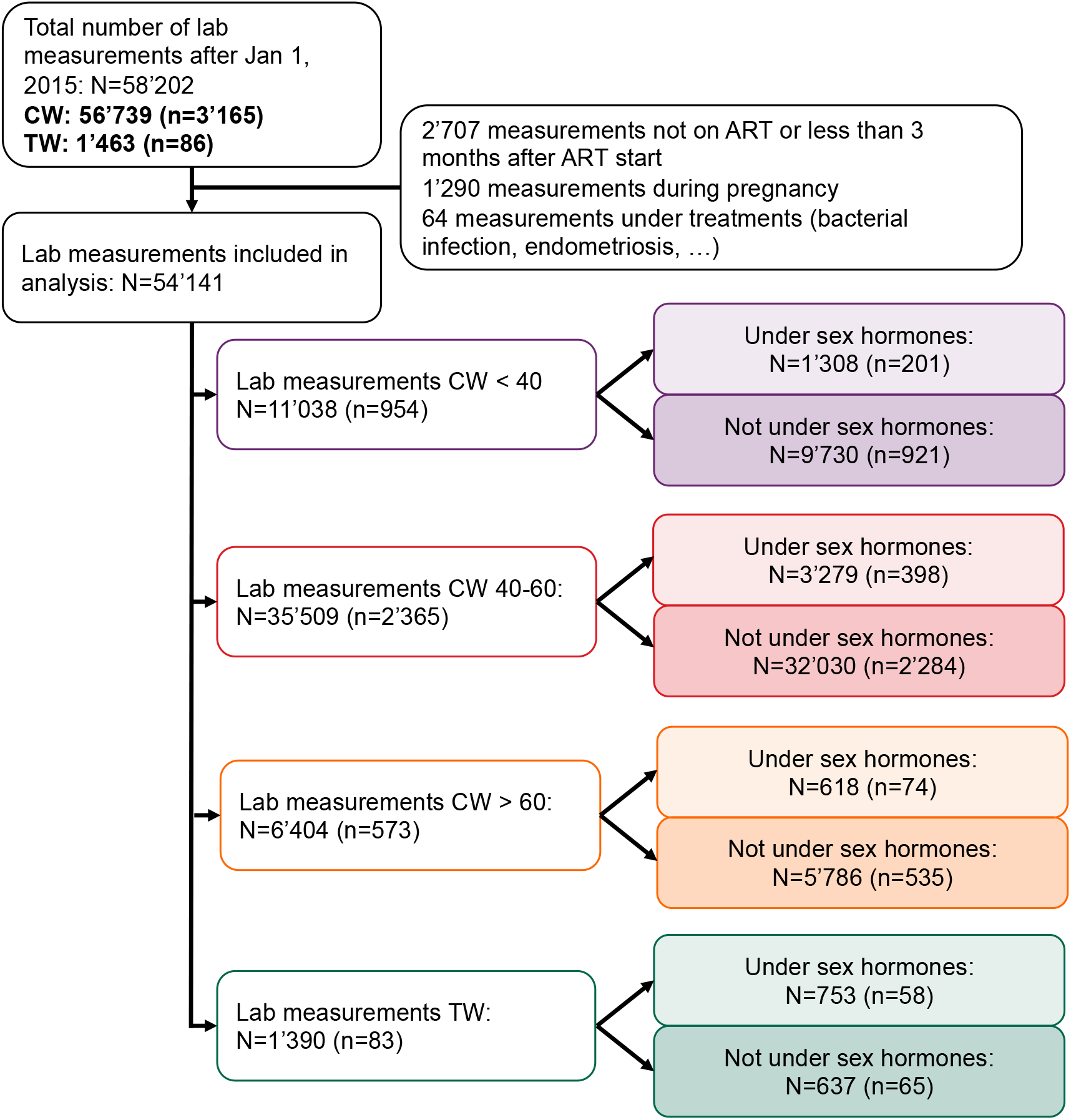
Flow chart of laboratory measurements selection and repartition in the different subgroups. N corresponds to the number of samples with lab measurements, n corresponds to the number of SHCS participants (one participant can have several samples with lab measurements included in the analysis and can contribute to both the no hormone intake and hormone intake groups).

As previously described (33), TW in the SHCS were more likely to be of Asian and Hispano-American ethnicity compared to CW and had overall similar education levels as CW (Table 1). A higher proportion of TW had ever taken sex hormones compared to CW (69.9% versus 18.9%). We observed the highest CD4+ T cell, CD8+ T cell, and lymphocyte counts in TW with sex hormone intake (median respectively 772 cells/μL, 822 cells/μL, and 2288 cells/μL), while the CD4:CD8 ratio was the lowest in TW without sex hormone intake (median 0.80) (Table 2, Figure 2, Supplementary Figure 1, 2). CW>60 with sex hormone intake had the lowest proportion (0.2%) of samples with detectable viral load (RNA > 50 copies/mL), and TW without sex hormone intake the highest (8.3%). Of note, only a small proportions of viral load measurements in TW without sex hormone intake were above 1’000 copies/mL (1.3%, Supplementary Table 1).

**Table 1:** Description of the population of cis women (CW) and trans women (TW) for which samples were included in the analysis

|  | CW | TW | Overall |
| --- | --- | --- | --- |
| **Sample size** | **n=3092** | **n=83** | **n=3175** |
| **Age in 2022:** |  |  |  |
| median (min,max) | 53 (18,95) | 45 (21,75) | 53 (18,95) |
| **Ethnicity: n(%)** |  |  |  |
| White | 1642 (53.1%) | 36 (43.4%) | 1678 (52.9%) |
| Black | 1103 (35.7%) | 5 (6.0%) | 1108 (34.9%) |
| Hispano-american | 104 (3.4%) | 16 (19.3%) | 120 (3.8%) |
| Asian | 227 (7.3%) | 25 (30.1%) | 252 (7.9%) |
| Other/unknown | 14 (0.5%) | 1 (1.2%) | 15 (0.5%) |
| Missing | 2 (0.1%) | 0 (0%) | 2 (0.1%) |
| **Education: n(%)** |  |  |  |
| Less than bachelor | 2329 (75.3%) | 63 (75.9%) | 2392 (75.3%) |
| Bachelor and more | 635 (20.5%) | 18 (21.7%) | 653 (20.6%) |
| Other/unknown | 47 (1.5%) | 2 (2.4%) | 49 (1.5%) |
| Missing | 81 (2.6%) | 0 (0%) | 81 (2.6%) |
| **IVD use: n(%)** |  |  |  |
| No | 2549 (82.4%) | 80 (96.4%) | 2629 (82.8%) |
| Yes | 518 (16.8%) | 3 (3.6%) | 521 (16.4%) |
| Missing | 25 (0.8%) | 0 (0%) | 25 (0.8%) |
| **Has taken sex hormones at least once since 2015: n(%)** |  |  |  |
| No | 2509 (81.1%) | 25 (30.1%) | 2534 (79.8%) |
| Yes | 583 (18.9%) | 58 (69.9%) | 641 (20.2%) |

**Table 2:** Description of lab measurements (CD4, CD8, and lymphocyte counts, CD4:CD8 ratio, detectable viral load) from the samples included in the statistical analysis. For each marker the number of measurements available (N) is displayed.

| Under sex hormone | CW < 40 |  | CW 40-60 |  | CW > 60 |  | TW |  | CM |
| --- | --- | --- | --- | --- | --- | --- | --- | --- | --- |
|  | No | Yes | No | Yes | No | Yes | No | Yes |  |
| **Total sample size (N)** | 9730 | 1308 | 32030 | 3279 | 5786 | 618 | 637 | 753 | 147298 |
| **Age at sample time:** | 36 (32,38) | 35 (30,38) | 51 (46,55) | 51 (45,55) | 66 (63,71) | 67 (62,76) | 45 (38,52) | 42 (37,50) | 52 (44,59) |
| **CD4 (cells/uL) (N)** | 8118 | 1108 | 27395 | 2805 | 5006 | 509 | 532 | 631 | 124823 |
| median (Q1,Q3) | 649 (480,861) | 715 (557,954) | 672 (488,887) | 720 (548,929) | 663 (477,878) | 720 (580,893) | 617 (426,892) | 772 (520,1006) | 651 (480,851) |
| **CD8 (cells/uL) (N)** | 8064 | 1103 | 27194 | 2793 | 5001 | 494 | 531 | 616 | 123685 |
| median (Q1,Q3) | 674 (507,899) | 656 (489,880) | 675 (491,913) | 651 (468,882) | 656 (459,900) | 557 (387,866) | 755 (583,975) | 822 (609,1033) | 732 (534,996) |
| **CD4:CD8 ratio (N)** | 8064 | 1103 | 27190 | 2793 | 5001 | 494 | 531 | 616 | 123679 |
| median (Q1,Q3) | 1.00 (0.70,1.35) | 1.16 (0.81,1.54) | 1.01 (0.71,1.39) | 1.14 (0.82,1.59) | 1.03 (0.67,1.50) | 1.25 (0.92,1.76) | 0.84 (0.56,1.12) | 0.94 (0.69,1.22) | 0.89 (0.62,1.24) |
| **Lymphocytes (cells/uL) (N)** | 7974 | 1096 | 26670 | 2663 | 4812 | 487 | 525 | 618 | 121563 |
| median (Q1,Q3) | 1873 (1500,2332) | 1908 (1529,2398) | 1900 (1500,2400) | 1937 (1569,2400) | 1900 (1476,2395) | 1870 (1558,2364) | 2082 (1700,2581) | 2288 (1768,2866) | 1949 (1548,2421) |
| **HIV-1 VL (copies/mL) (N)** | 9586 | 1288 | 31270 | 3212 | 5689 | 612 | 612 | 745 | 144228 |
| Detectable VL (>50): n(%) | 698 (7.2%) | 45 (3.4%) | 1499 (4.7%) | 92 (2.9%) | 256 (4.4%) | 1 (0.2%) | 53 (8.3%) | 12 (1.6%) | 6078 (4.1%) |

**Figure 2:**
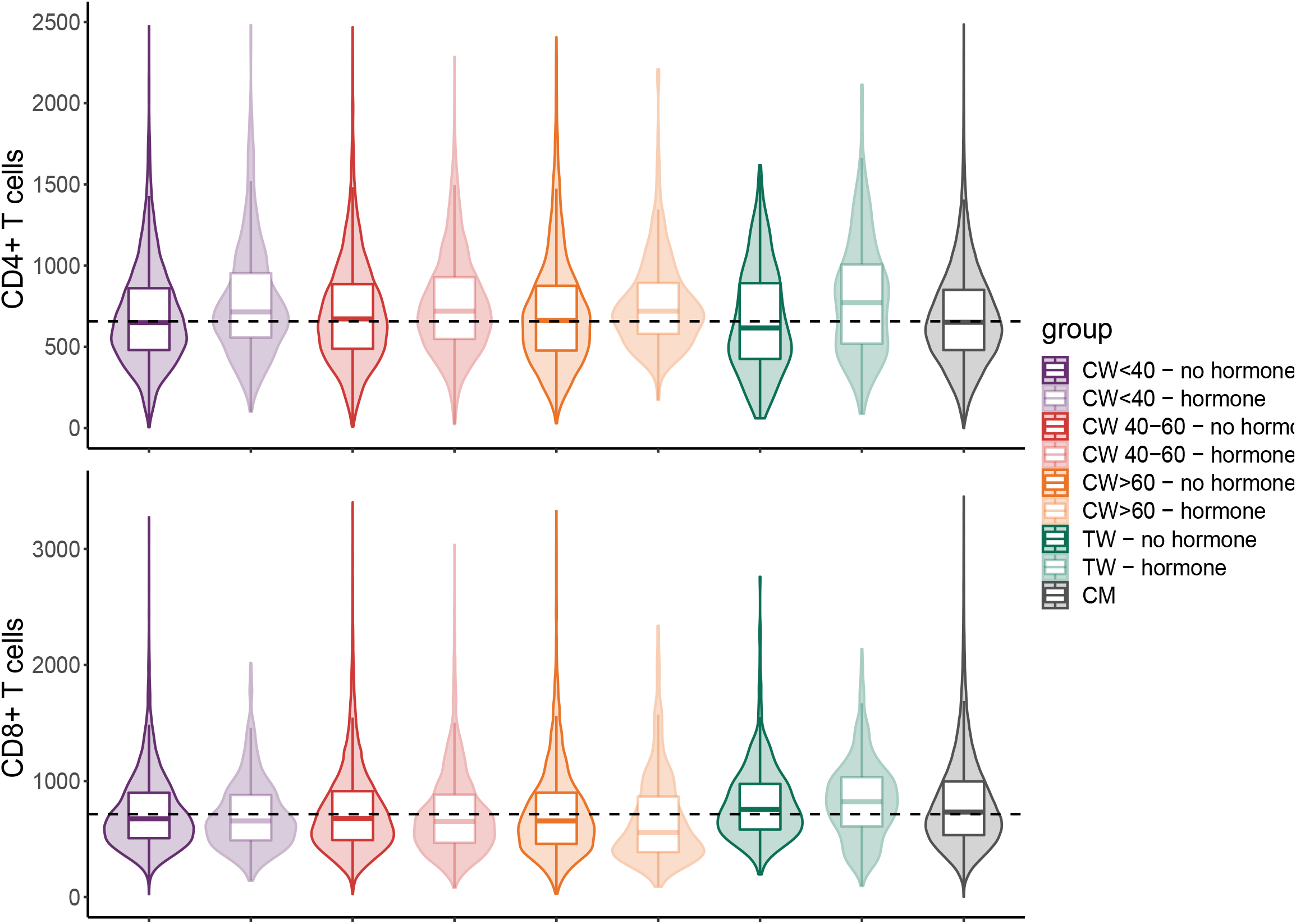
Distribution of CD4+ and CD8+ T cell counts in the different studied populations: CW aged 40 and younger (purple), CW 40-60 years-old (red), CW older than 60 years-old (orange), TW (green) and cis men (CM, black). Dashed line corresponds to the median among all measurements (CD4=657 cells/μL, CD8=715 cells/μL). For display purposes, 34 measurements (CD4>2500 cells/μL) and 77 measurements (CD8>3500 cells/μL) were excluded from the plot. Individual data points are shown in addition in Supplementary Figure 2.

### Diversity of sex hormone intake in women from the Swiss HIV Cohort Study

Using information contained in the ATC codes, we were able to further differentiate the type of sex hormones taken by women from the SHCS, as well as their route of administration (Table 3, Supplementary Figure 3). CW<40 mostly reported the use of systemic progestogens (35.1%) or combined oestrogens and progestogens (36.9%), in line with the use of hormonal contraception (HC). CW>60 mostly reported the use of local oestrogens (63.2%) or systemic combined oestrogens and progestogens (21.1%), corresponding to the use of menopause hormone therapy (MHT). In the group of CW 40-60, we observed more heterogeneity, with reports of HC use (systemic combined oestrogens and progestogens, 38.4%, and systemic progestogens, 21.1%) but also MHT (systemic and local oestrogens, respectively 17.6% and 14.5%), reflecting the potential overlap of contraception with the need for managing symptoms during perimenopause. Finally, TW mostly reported the use of oestrogens (53.2%) or combined oestrogens and antiandrogens (32.9%). Our descriptive analysis highlighted the diversity of sex hormone intake in women from the SHCS and the complexity of the data when assessing and comparing different age groups.

**Table 3:**
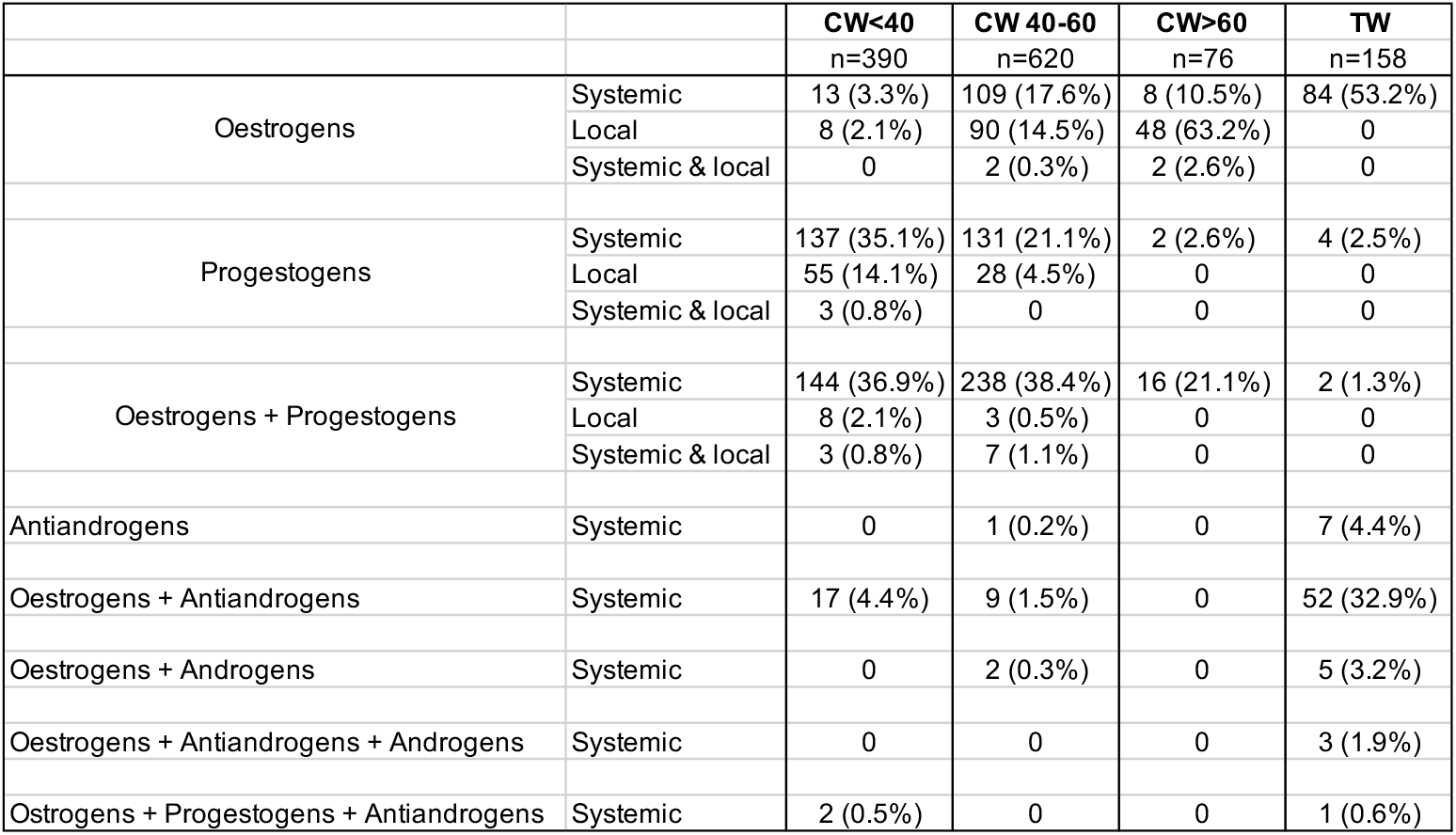
Description of sex hormones intake reported by women in the SHCS. Sex hormone reports are defined as explained in the supplementary figure 3. Total number of sex hormone reports in each group are displayed (n).

|  |  | CW<40<br>n=390 | CW 40-60<br>n=620 | CW>60<br>n=76 | TW<br>n=158 |
| --- | --- | --- | --- | --- | --- |
| Oestrogens | Systemic | 13 (3.3%) | 109 (17.6%) | 8 (10.5%) | 84 (53.2%) |
|  | Local | 8 (2.1%) | 90 (14.5%) | 48 (63.2%) | 0 |
|  | Systemic & local | 0 | 2 (0.3%) | 2 (2.6%) | 0 |
| Progestogens | Systemic | 137 (35.1%) | 131 (21.1%) | 2 (2.6%) | 4 (2.5%) |
|  | Local | 55 (14.1%) | 28 (4.5%) | 0 | 0 |
|  | Systemic & local | 3 (0.8%) | 0 | 0 | 0 |
| Oestrogens + Progestogens | Systemic | 144 (36.9%) | 238 (38.4%) | 16 (21.1%) | 2 (1.3%) |
|  | Local | 8 (2.1%) | 3 (0.5%) | 0 | 0 |
|  | Systemic & local | 3 (0.8%) | 7 (1.1%) | 0 | 0 |
| Antiandrogens | Systemic | 0 | 1 (0.2%) | 0 | 7 (4.4%) |
| Oestrogens + Antiandrogens | Systemic | 17 (4.4%) | 9 (1.5%) | 0 | 52 (32.9%) |
| Oestrogens + Androgens | Systemic | 0 | 2 (0.3%) | 0 | 5 (3.2%) |
| Oestrogens + Antiandrogens + Androgens | Systemic | 0 | 0 | 0 | 3 (1.9%) |
| Oestrogens + Progestogens + Antiandrogens | Systemic | 2 (0.5%) | 0 | 0 | 1 (0.6%) |

### Sex hormone intake is associated with distinct effects on immune markers in CW and TW with HIV

We assessed the effects of sex hormone intake on immune markers by using linear mixed-effect models adjusted for potential confounders such as years since ART start, ethnicity, intravenous drug use, education level, and age (Figure 3, Supplementary Figure 4). We found significantly different effects of sex hormone intake on CD4 and CD4:CD8 ratio between the studied groups (CW<40, CW 40-60, CW>60, and TW) (interaction p^CD4^=0.02, p^ratio^=0.007). Interestingly, we found a significant increase of CD4 counts, CD4:CD8 ratio, and lymphocyte counts associated with sex hormone intake in TW (effect on the scaled outcomes = 0.19, 95% confidence interval [0.10,0.28] for CD4 counts, 0.08 [0.02,0.15] for CD4:CD8 ratio, and 0.15 [0.04,0.25] for lymphocytes counts). CD4 and lymphocyte counts displayed high correlation in TW (Pearson r=0.78, p<0.001, Supplementary Figure 5), suggesting that the increase in lymphocyte counts in TW with sex hormone intake was mostly due to the observed increase in CD4 counts.

**Figure 3:**
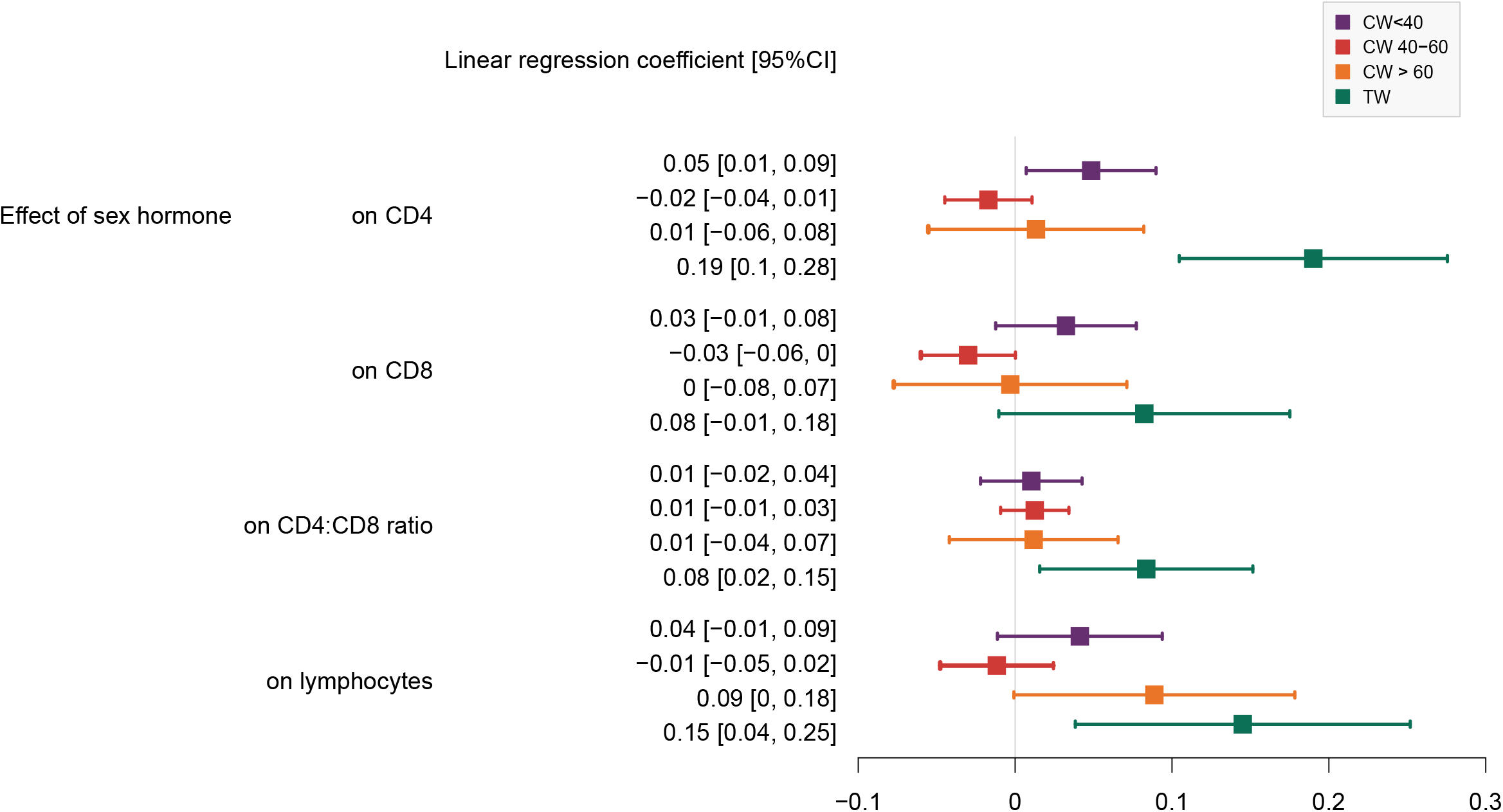
Effect of sex hormone intake in CW<40 (purple), CW 40-60 (red), CW>60 (orange), and TW (green) on the scaled studied outcomes: CD4 counts, CD8 counts, CD4:CD8 ratio and lymphocytes counts. Models were adjusted for potential confounders such as years since first start of ART, ethnicity, intravenous drug use, education level, and age.

Chronic viral and bacterial infections and lifestyle factors such as smoking could have an impact on immune markers (41-43). We conducted sensitivity analyses adjusted on smoking status and co-infection with hepatitis C and/or syphilis and found consistent results on the effect of sex hormones on immune markers (Supplementary Figure 6). We further investigated whether changes in immune markers could be linked to viral control, adherence to ART, and/or mental health problems. Interestingly, sex hormone intake was significantly correlated with undetectable or suppressed viral load in both CW and TW (chi square test p<0.001 for CW<40, CW>60, and TW, and p=0002 for CW 40-60, Supplementary Table 1), with better self-reported adherence in CW<40 (chi square test p<0001) and CW 40-60 (chi square test p<0.001), but with more depression in CW<40 (chi square test p=0.025). When adjusting for depression, self-reported adherence, and viral load, the increase of CD4 counts in TW upon sex hormone intake remained significant (effect on the scaled outcome = 0.14 [0.05,0.23], Supplementary Figure 7), suggesting that this increase cannot solely be explained by better adherence to treatment and control of the virus.

We further investigated whether the effects of sex hormones on immune markers in CW and TW were comparable to the differences in immune marker levels between cis men and CW (Supplementary Figure 8). Interestingly, the size of the effect of sex hormones on CD4 counts in TW was very similar to the difference of CD4 counts in CW (without sex hormone intake) versus cis men (CM, effect on the scaled outcome = 0.19 [0.14,0.24]). However, the effect on CD8 counts and lymphocytes was different, suggesting that sex and gender differences in immune markers cannot fully be compared to the inferred effect of exogenous sex hormone intake in CW and TW.

Considering the diversity of sex hormones taken by the population of women in the SHCS, we also investigated the effect of different type and route of hormones among the defined groups (Figure 4). Among TW taking sex hormones, we found trends for lower CD4, CD8, and lymphocyte counts in TW with combinations of oestrogens and antiandrogens versus TW taking oestrogens only (p^CD4^=0.075, p^CD8^=0.052, and p^lymphocytes^=0.090). Among CW<40 taking sex hormones, we observed significantly lower CD8 and lymphocyte counts and higher CD4:CD8 ratio (p^CD8^=0.050, p^lymphocyte^=0.015, p^ratio^=0.004 respectively) among those taking progestogens only versus those taking oestrogens and progestogens in combination.

**Figure 4:**
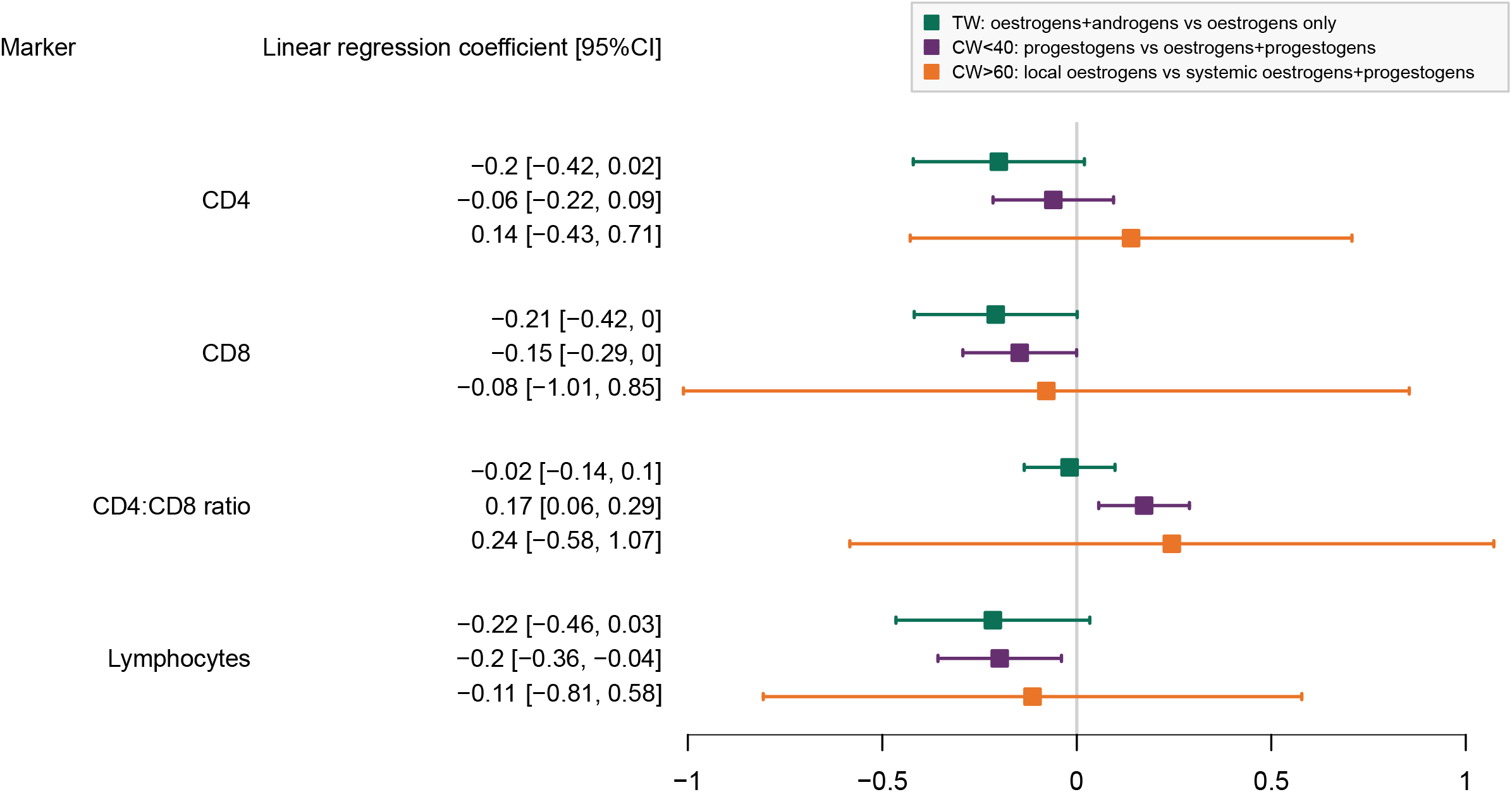
Effect of different type and route of hormones on immune markers: in green, comparison of TW taking oestrogens + androgens versus oestrogens only; in purple, comparison of CW<40 taking progestogens versus oestrogens+progestogens; in orange, comparison of CW>60 taking local oestrogens versus systemic oestrogen+progestogens. Models were adjusted for potential confounders such as years since first start of ART, ethnicity, intravenous drug use, education level, and age.

### No evidence for inflammatory pathways activation upon sex hormone intake in TW

We further sought to identify whether sex hormone intake had an anti-inflammatory effect in TW (44). To do so, we assessed the effect of sex hormone intake on immunity in 31 TW (19 taking oestrogens only and 12 taking a combination of oestrogens and antiandrogens) by undertaking a targeted proteomic analysis of individual soluble inflammation-associated proteins in two sequential samples: one taken up to 2 years before the start of sex hormone intake (median 176 days, 1Q-3Q = 58-240 days) and one taken between 1 and 18 months after the start of sex hormone intake (median 175 days, 1Q-3Q = 155-225 days). Using an unsupervised hierarchical clustering strategy, we found no distinct immune signature between the samples taken before and after sex hormone intake (Supplementary Figure 9). After adjustment for multiple testing, we found no significant differential protein concentration in TW before and after sex hormone intake (Supplementary Table 2). Only two proteins displayed significant changes when considering the p-value unadjusted for multiple testing, namely neurotrophin-3 (NT-3, p=0.019) and STAMBP (p=0.032), for which we observed a decreased concentration upon sex hormone intake (Supplementary Figure 10).

## Discussion

We report here on the first large cohort study analyzing the impact of exogenous sex hormone intake on the immune system in women with HIV. Our study explicitly included TW in the analysis to gain insights into effects of sex hormone intake on the immune system in a population often understudied in HIV-1 research.

Although rates of MHT intake in menopausal cis women were previously reported in the SHCS (45), we were now able to extensively describe the diversity of sex hormones taken by women in the SHCS. Notably, our analysis highlighted the complexity of the menopausal transition, during which the need of HC and MHT are likely to overlap.

Based on a number of studies partially attributing sex and gender differences in immunity to the role of sex hormones (1-5, 16), we investigated how exogenous sex hormone intake affect major HIV-1 immune markers. Observed differences of sex hormone intake on immune markers (CD4, CD4:CD8 ratio) in CW and TW could be due to differences in the combination of hormones taken. Indeed, TW mostly reported intake of oestrogens and/or antiandrogens and CW mostly reported intake of oestrogens and/or progestogens, depending on their age. Additionally, within each group, we did observe significant (in CW<40) or trends (in TW) of differential effects on HIV-1 markers by type of sex hormone.

Differences in dosing and mechanisms of actions of sex hormones on the immune system in CW and TW could also explain our results and should be further investigated.

Our most robust result was the significant association between sex hormone intake and an increase in CD4 counts in TW. This increase was similar in magnitude to the difference of CD4 counts between CW (not taking sex hormones) and CM, but this similarity was not identified for the other markers (CD8, CD4:CD8 ratio, lymphocytes). In line with previous work (46-48), this highlights the complexity and variety of factors (chromosomal, hormonal, and social) playing a role in immune modulation.

Additionally, we explored how factors such as adherence and mental health could play a role in our analyses. Indeed, it has been reported that sex hormone intake in TW can be associated with lower adherence to ART, driven by concerns of drug-drug interactions (38, 39, 49). However, access to gender-affirming hormone therapy in TW was also shown to be associated with overall better mental health (50-52), which is known to be a good predictor for adherence in people with HIV (53-56). In our analysis, we did not find any association in TW between sex hormone intake and self-reported adherence to ART or depression. Even after adjusting for these variables, we still found a significant increase in CD4 counts with sex hormone intake in TW, suggesting that other social and biological factors are potentially responsible for this increase. However, we did identify a correlation between depression and HC in CW<40, in line with previous reports (57-59). Although previous results showed that mental health was not associated with adherence to ART or viral suppression during menopausal transition (60), more studies are needed in CW and TW with HIV to investigate the interplay between sex hormone intake, mental health, and immunity.

Using a high-throughput serum proteomics approach, we aimed at identifying the inflammatory pathways potentially modulated with sex hormone intake in TW. We could not identify any proteins with significant concentration change before and after sex hormone intake in TW after adjusting for multiple testing. This negative result could be due to our low sample size or to the timing of the sampling before and after the start of sex hormone intake. Indeed, a previous report found that inflammatory markers were only transiently modified in TW receiving oral oestrogens, with a return to baseline levels between 2 to 6 months after the start of oestrogens intake (61). As the samples retrieved from our biobank were obtained with a median of around 6 months after the start of sex hormone intake, we cannot exclude that this time window was not allowing us to identify transient inflammatory changes happening earlier after the start of sex hormone intake. Although all our statistical analyses were adjusted for confounders and we conducted several sensitivity analyses, this study is observational by design, and it is therefore not possible to definitely conclude on the causality between sex hormone intake and increase of CD4+ T cell counts in TW. Our analysis did not include longitudinal modeling of immune trajectories, preventing us to draw final conclusions on the time between start of sex hormone intake and potential modulations of immune markers. However, measurements at more regular and specific timepoints would be required to address this question. Additionally, we only observed the total number of cells, and the cohort design does not include any phenotyping of cells, which is preventing us to conclude on the cell subtypes with increased population with sex hormone intake.

In conclusion, our results present new insights on the effect of exogenous sex hormone intake on the immune system of cis and trans women with HIV and highlights the need to consider the potential role of sex hormones in modulating the immune system among other biological and social factors.

## Methods

### Swiss HIV Cohort Study

The Swiss HIV Cohort Study (SHCS) is a prospective multicenter cohort study enrolling people with HIV in Switzerland (62). The SHCS was approved by the local ethical committees of the participating centers, and written informed consent was obtained from all participants. Participants are followed-up biannually with data collection including behavioral information, drug intake, gynecological exam if applicable. Blood samples are systematically drawn at each visit for laboratory measurements and storage in a biobank. In the main analysis, we considered laboratory measurements from samples drawn in CW and TW after January 1^st^, 2015, when systemic report of most comedications taken by SHCS participants started. Reports include (since 2015) ATC codes, drug brand, dose, and route of administration for both antiretrovirals and co-medication. We excluded from the analysis laboratory measurements that were sampled when participants were not on ART, less than 3 months after the first start of ART intake, during pregnancies, and during hormone intake as subsequent treatment for bacterial vaginosis or treatment for endometriosis (as these conditions could themselves be associated with changes in immunity).

As a comparison we also considered laboratory measurements in 147’298 samples on ART for at least 3 months from 8’611 cis men after January 1^st^, 2015.

### Cis and trans women in the SHCS

We identified 86 TW – using criteria previously described (33), and 3’165 CW with at least one laboratory measurement after January 1^st^, 2015.

### Sex hormone intake definition

Sex hormone intake was assessed using the following ATC codes: G02BA03 (hormonal intrauterine contraceptive with progestogens), G02BB (contraceptive vaginal ring with oestrogens and progestogens in combination), G03A (hormonal contraceptive for systemic use), G03C (oestrogens), G03D (progestogens), G03E (androgens and female sex hormone in combination), and G03F (progestogens and oestrogens in combination). For each ATC code, report of the start and end of the corresponding drug are indicated. Sex hormone reports for each participant were obtained by combining different sex hormones when taken at the same time (Supplementary Figure 3), then classified by the type (e.g., oestrogens, progestogens, oestrogens + antiandrogens) and route (e.g., systemic, local) of intake. For example, a participant’s report with a simultaneous intake of a product with ATC code G03CA (transdermal, oestrogens) and another product with ATC code G03AC (systemic, progestogens) will be classified as systemic oestrogens + progestogens.

Measurements that were realized on blood samples drawn during the period of sex hormone intake were labeled as “with sex hormone intake”, others “without sex hormone intake”.

### Smoking, depression, and adherence definition

Information on smoking, depression, and self-reported adherence to ART (obtained during bi-annual visits) were linked to laboratory measurements realized up to 6 months before the corresponding follow-up visit.

### Proteomics measurements

We retrieved plasma samples from the SHCS biobank: we selected 31 TW for which plasma samples were available both before the first reported start of sex hormone intake (between one month and two years) and after it (between three months and two years), with no reported interruption between the start of intake and the plasma collection. Proteomics measurements were realized on the 62 retrieved samples using the Olink® Target 96 technology Inflammation panel (63), which enables high-throughput, multiplex immunoassays by measuring up to 92 proteins across 96 plasma samples simultaneously (Olink Bioscience AB, Uppsala, Sweden). Olink® Target 96 is based on the Proximity Extension Assay (PEA). Using oligonucleotide-labeled antibody probe pairs, PEA permits simultaneous assessment of multiple proteins, maintaining the precision of dual epitope ELISA without the loss of specificity of earlier generation multiplex assays. Signal production requires both recognition of DNA barcoding from sequence-specific oligonucleotides and dual recognition of correctly matched antibody pairs. The PEA assay reports fold change in log_2_ (Normalized Protein eXpression [NPX]) units. The Inflammation panel informs on factors involved in cell differentiation and cytokine-mediated signaling pathways.

### Statistical analysis

Studied outcomes (CD4+ T cell count, CD8+ T cell count, CD4:CD8 ratio, and total lymphocyte count) were transformed using square-root transformation for the cell counts and log transformation for the ratio, and then scaled. Each outcome was then expressed using a linear mixed effect model with fixed effects for the adjustment variable and a random effect for the individuals to account for repeated measurements. The linear mixed effect model can be written as follows:

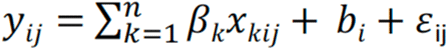

Where y_ij_ is the j^th^ observation of participant i, x_k_ is the k^th^ of n fixed effects (variable of adjustment) and β_k_ its corresponding coefficient, b_i_ is the participant-specific normally distributed random effect and ε_ij_ the normally distributed residual error.

For the fixed effect, we included a “group” variable (CW<40, CW 40-60, CW>60, TW) and a variable “with sex hormone intake” (yes, no). We considered CW in different age groups due to the strong correlation between the type of sex hormones used and the age of CW (i.e., CW<40 mostly take hormonal contraception, CW>60 mostly take hormonal replacement therapy, while the hormones taken by CW 40-60 are more heterogeneous). We tested whether the sex hormone intake was associated to the outcome differentially in the respective groups by estimating an interaction between the group variable and the sex hormone intake variable. The reference category was CW 40-60, as this was the group with the biggest sample size. We assessed the significance of the interaction term using a likelihood ratio test (complete model with interaction term, reduced model without interaction term). We report the effect of sex hormone intake in the different groups on the scaled outcomes and p-value of the likelihood ratio test for the interaction term. To control for potential confounders, we adjusted the models on the following variables: age at time of sample (continuous with splines), ethnicity, education level, time since ART start (continuous with splines), use of intravenous drugs. In sensitivity analyses, we also adjusted for depression, smoking status at time of sampling, self-reported adherence (three categories: never missed, missed once a month, and missed more than once every two weeks) and viral load (three categories: below 50 copies/mL, 50-1’000 copies/mL and >1’000 copies/mL).

Given the left-censored nature of the proteomics data, the analysis was conducted in two steps: we first pre-selected proteins of interest, by excluding those with a proportion of censored values among the 62 samples more than 75%, and a dispersion value (standard deviation divided by the mean value) less than 5%. We then tested for differences between pre and post sex hormone intake samples using paired t-tests. Paired t-tests in proteins with left-censored data were realized using the function cen_paired from the NADA2 R package. These analyses were conducted on all samples, on the subset of samples in participants taking oestrogens only, and on the subset of samples in participants taking a combination of oestrogens and anti-androgens. Multiple testing was corrected using the FDR method.

## Supporting information

All supplementary files

## Data Availability

The individual level datasets generated or analyzed during the current study do not fulfill the requirements for open data access:
1) The SHCS informed consent states that sharing data outside the SHCS network is only permitted for specific studies on HIV infection and its complications, and to researchers who have signed an agreement detailing the use of the data and biological samples; and
2) the data is too dense and comprehensive to preserve patient privacy in persons living with HIV.
According to the Swiss law, data cannot be shared if data subjects have not agreed or data is too sensitive to share. Investigators with a request for selected data should send a proposal to the respective SHCS address (www.shcs.ch/contact). The provision of data will be considered by the Scientific Board of the SHCS and the study team and is subject to Swiss legal and ethical regulations, and is outlined in a material and data transfer agreement.

## Acknowledgments

We thank the participants of the SHCS, the physicians and study nurses for excellent patient care, and the SHCS data center for excellent data management.

## Members of the Swiss HIV Cohort Study

Abela I, Aebi-Popp K, Anagnostopoulos A, Battegay M, Bernasconi E, Braun DL, Bucher HC, Calmy A, Cavassini M, Ciuffi A, Dollenmaier G, Egger M, Elzi L, Fehr J, Fellay J, Furrer H, Fux CA, Günthard HF (President of the SHCS), Hachfeld A, Haerry D (deputy of “Positive Council”), Hasse B, Hirsch HH, Hoffmann M, Hösli I, Huber M, Jackson-Perry D (patient representatives), Kahlert CR (Chairman of the Mother & Child Substudy), Kaiser L, Keiser O, Klimkait T, Kouyos RD, Kovari H, Kusejko K (Head of Data Centre), Labhardt N, Leuzinger K, Martinez de Tejada B, Marzolini C, Metzner KJ, Müller N, Nemeth J, Nicca D, Notter J, Paioni P, Pantaleo G, Perreau M, Rauch A (Chairman of the Scientific Board), Salazar-Vizcaya L, Schmid P, Speck R, Stöckle M (Chairman of the Clinical and Laboratory Committee), Tarr P, Trkola A, Wandeler G, Weisser M, Yerly S.

## Authors contribution

CP, RDK: conception and design of the study, analysis and interpretation of data, drafting and revision of the article. IAA: conception and design of the study, acquisition of data, analysis and interpretation of data, drafting and revision of the article. KAP: acquisition of data, analysis and interpretation of data, drafting and revision of the article. All other co-authors: acquisition of data, drafting and revision of the article.

## Data availability

The individual level datasets generated or analyzed during the current study do not fulfill the requirements for open data access:

1. The SHCS informed consent states that sharing data outside the SHCS network is only permitted for specific studies on HIV infection and its complications, and to researchers who have signed an agreement detailing the use of the data and biological samples; and
2. the data is too dense and comprehensive to preserve patient privacy in persons living with HIV.

According to the Swiss law, data cannot be shared if data subjects have not agreed or data is too sensitive to share. Investigators with a request for selected data should send a proposal to the respective SHCS address (www.shcs.ch/contact). The provision of data will be considered by the Scientific Board of the SHCS and the study team and is subject to Swiss legal and ethical regulations, and is outlined in a material and data transfer agreement.

