## Supplementary material for "Delineating the effect of sex hormone intake on immunity in cis and trans women with HIV": All supplementary files

**Supplementary Figure 1.** Distribution of CD4:CD8 ratio and lymphocyte counts in the different studied populations: CW aged 40 and younger (purple), CW 40-60 years-old (red), CW older than 60 years-old (orange), TW (green) and cis men (CM, black). Dashed line corresponds to the median among all measurements (ratio=0.93, lymphocytes=1938 cells/μL). For display purposes, 34 measurements (ratio>8) and 61 measurements (lymphocytes>7000 cells/μL) were excluded from the plot.

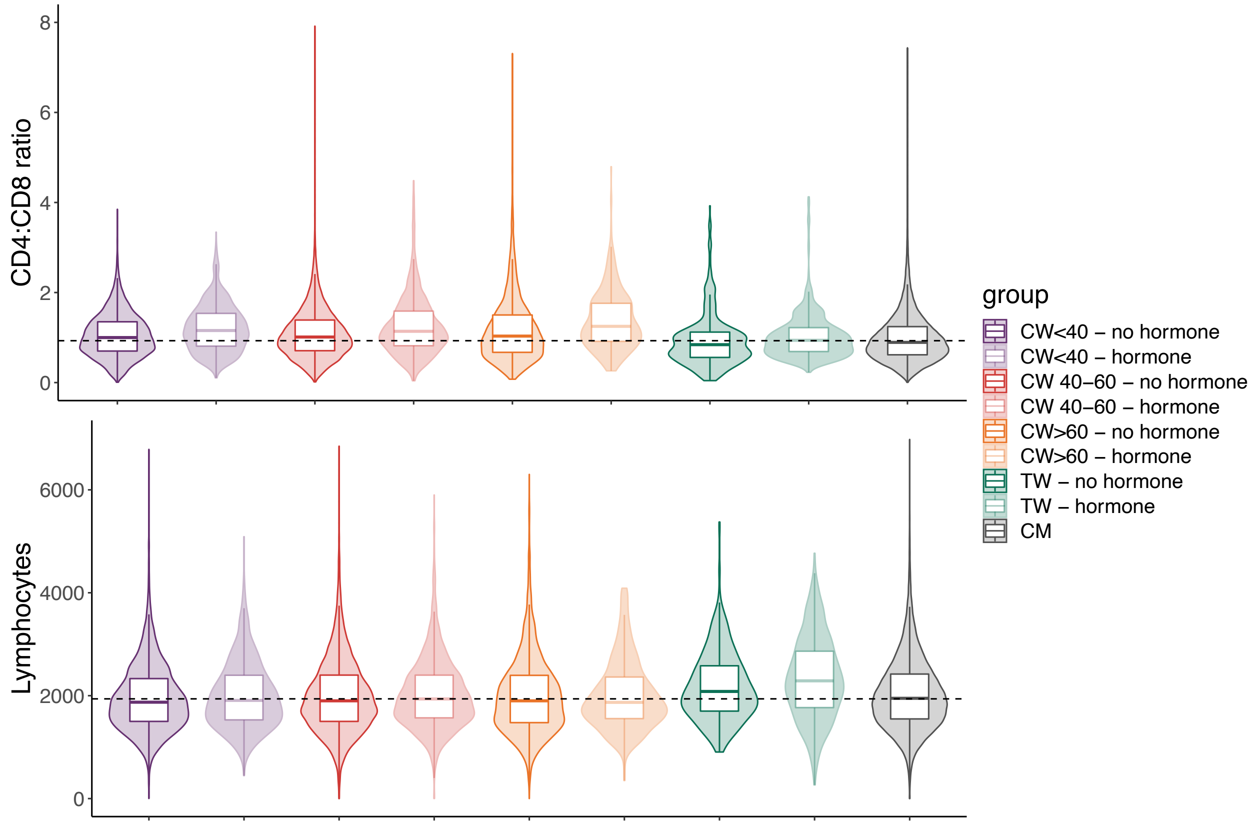

**Supplementary Figure 2:** Distribution of CD4+ and CD8+ T cell counts in the different studied populations: CW aged 40 and younger (purple), CW 40-60 years-old (red), CW older than 60 years-old (orange), TW (green) and cis men (CM, black). Dashed line corresponds to the median among all measurements (CD4=657 cells/μL, CD8=715 cells/μL). For display purposes, 34 measurements (CD4>2500 cells/μL) and 77 measurements (CD8>3500 cells/μL) were excluded from the plot. Each dot corresponds to an individual laboratory measurement.

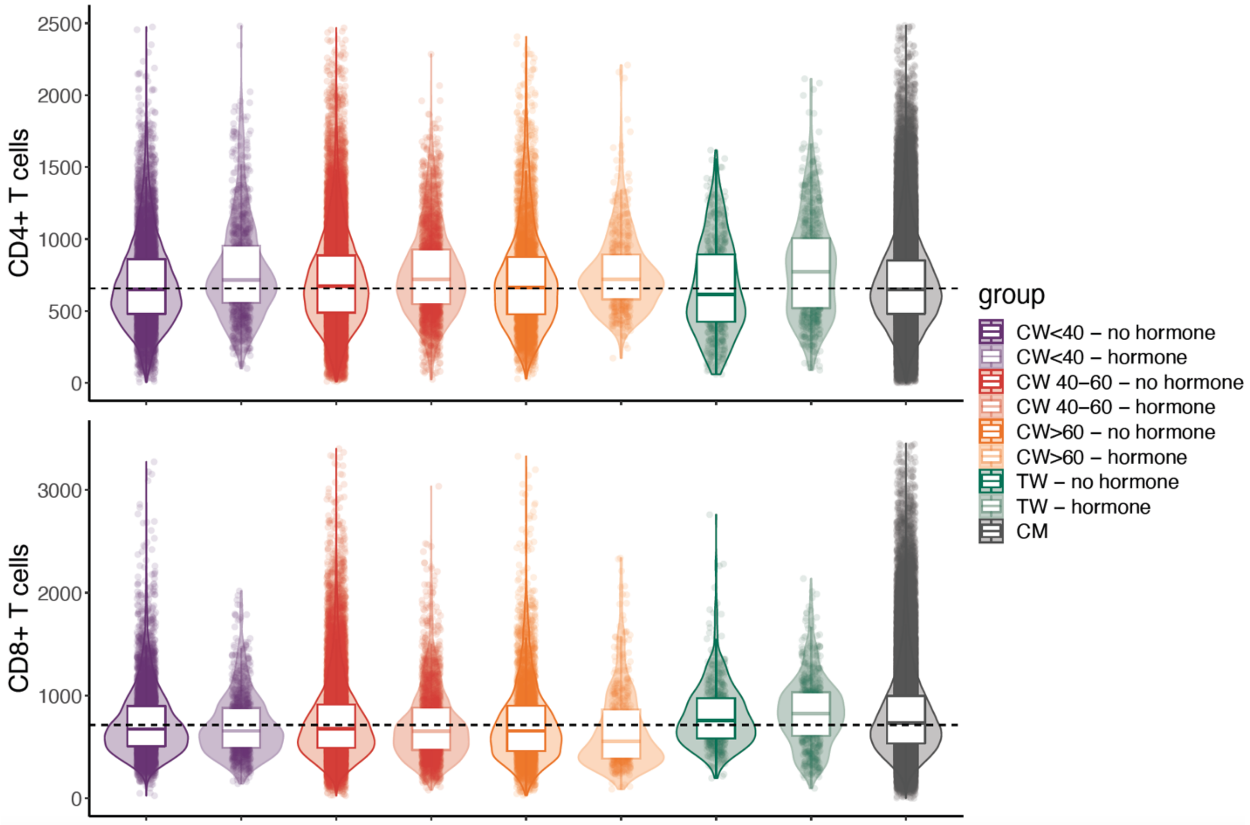

**Supplementary Figure 3.** Method of defining sex hormone reports using data from the SHCS. Products are combined when they overlap on a given period of time. Each period starting with a new product or combination of products is considered as a new report.

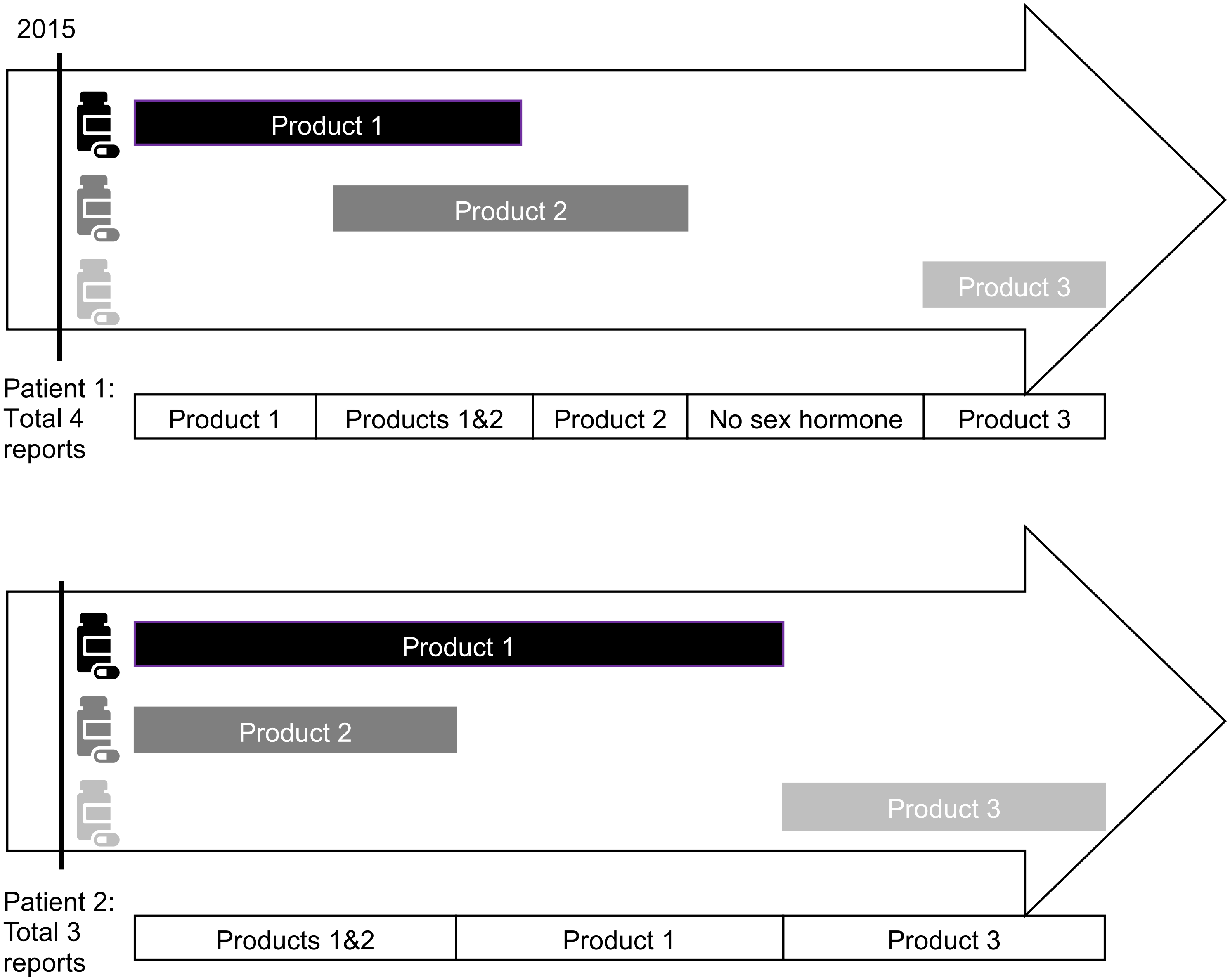

**Supplementary Figure 4.** Effect of sex hormone intake in CW<40 (purple), CW 40-60 (red), CW>60 (orange), and TW (green) on the scaled studied outcomes: CD4 counts, CD8 counts, CD4:CD8 ratio and lymphocytes counts in univariable models (i.e., unadjusted for confounders).

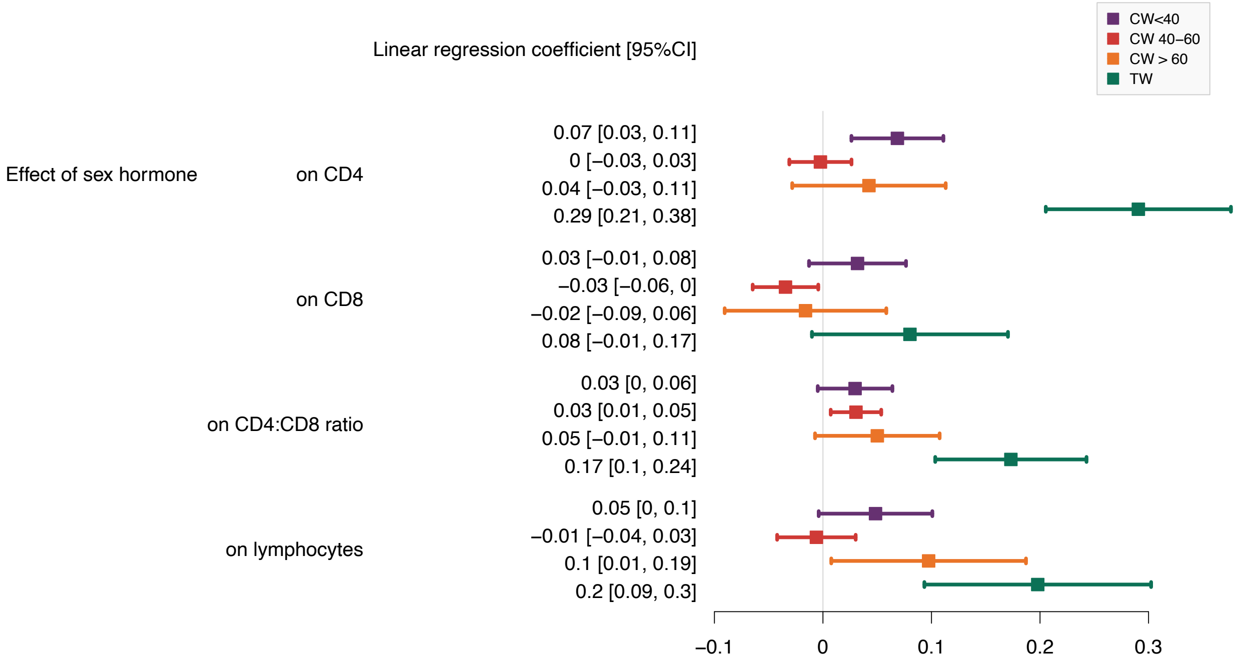

**Supplementary Figure 5.** Lymphocyte counts versus CD4 counts in TW not under sex hormone (dark green) and under sex hormone (light green)

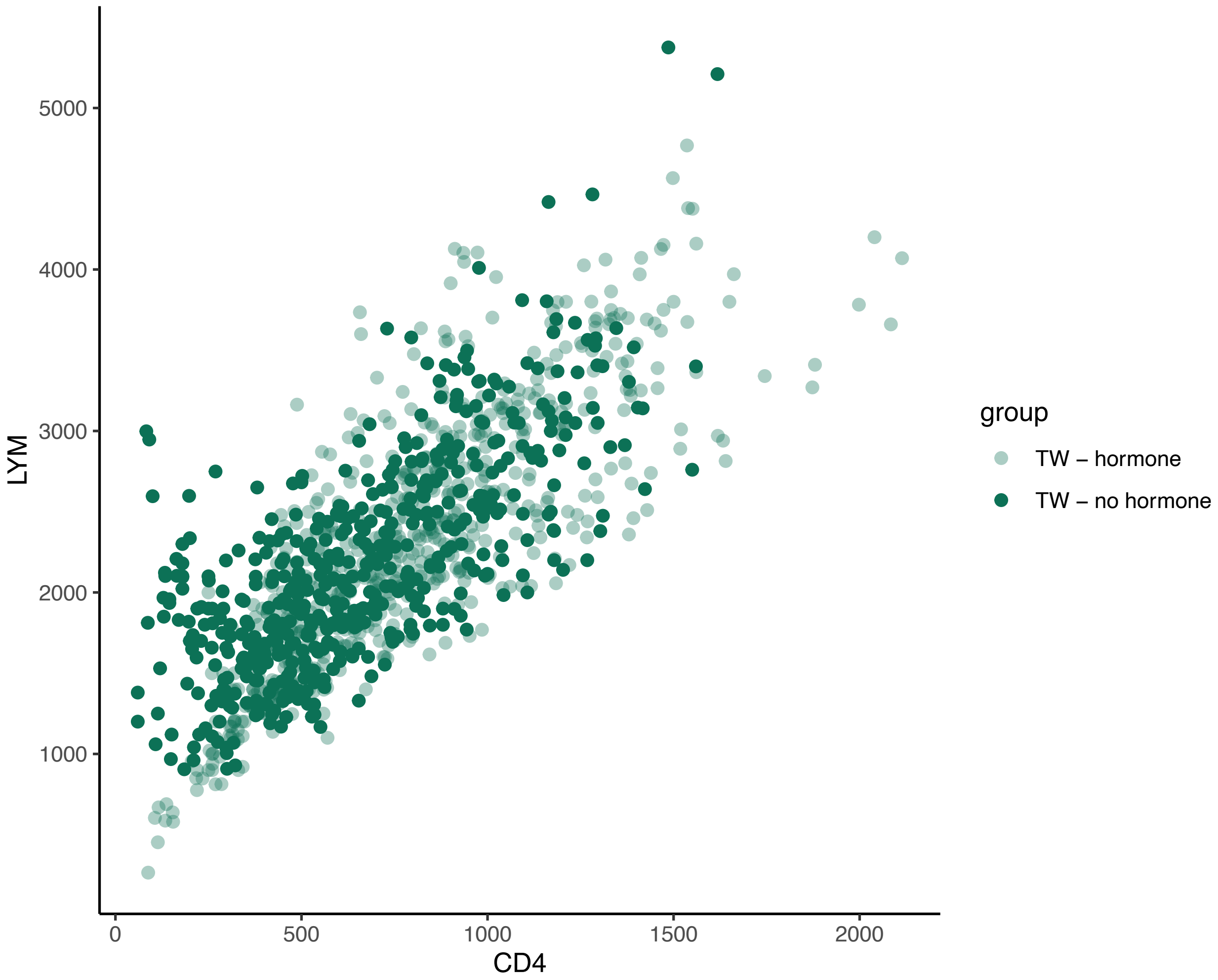

**Supplementary Figure 6.** Sensitivity analysis adjusted on smoking and co-infections.

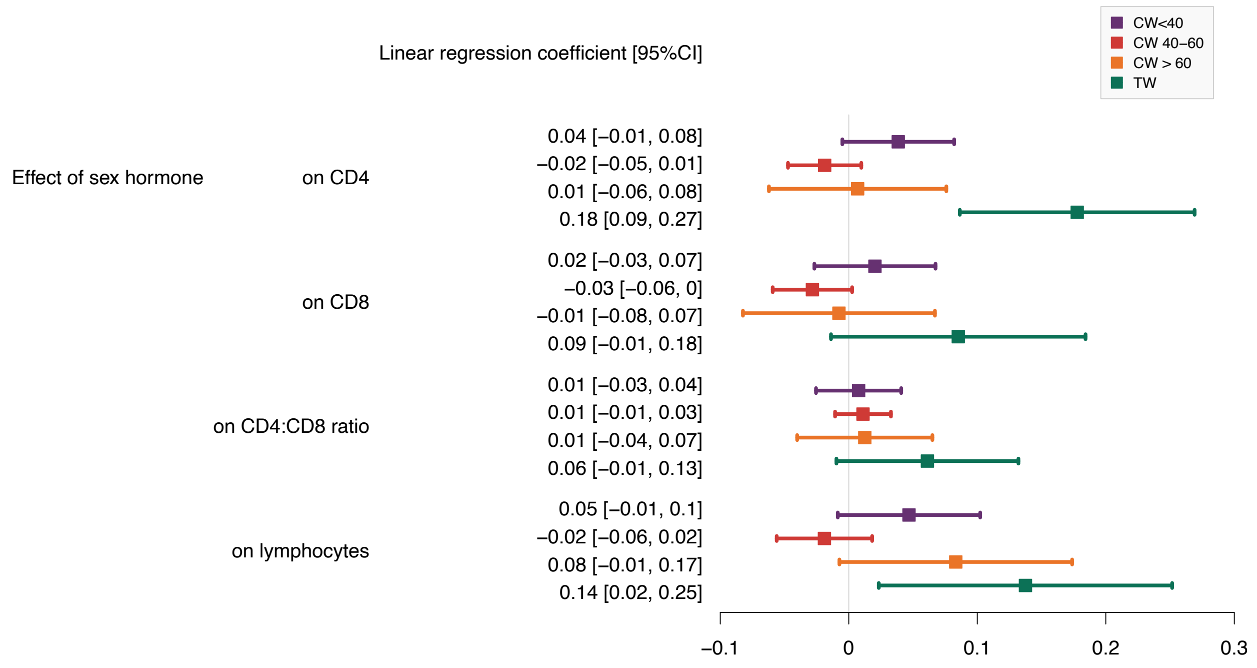


**Supplementary Figure 7.** Sensitivity analysis adjusted on viral load, self-reported adherence, and depression.

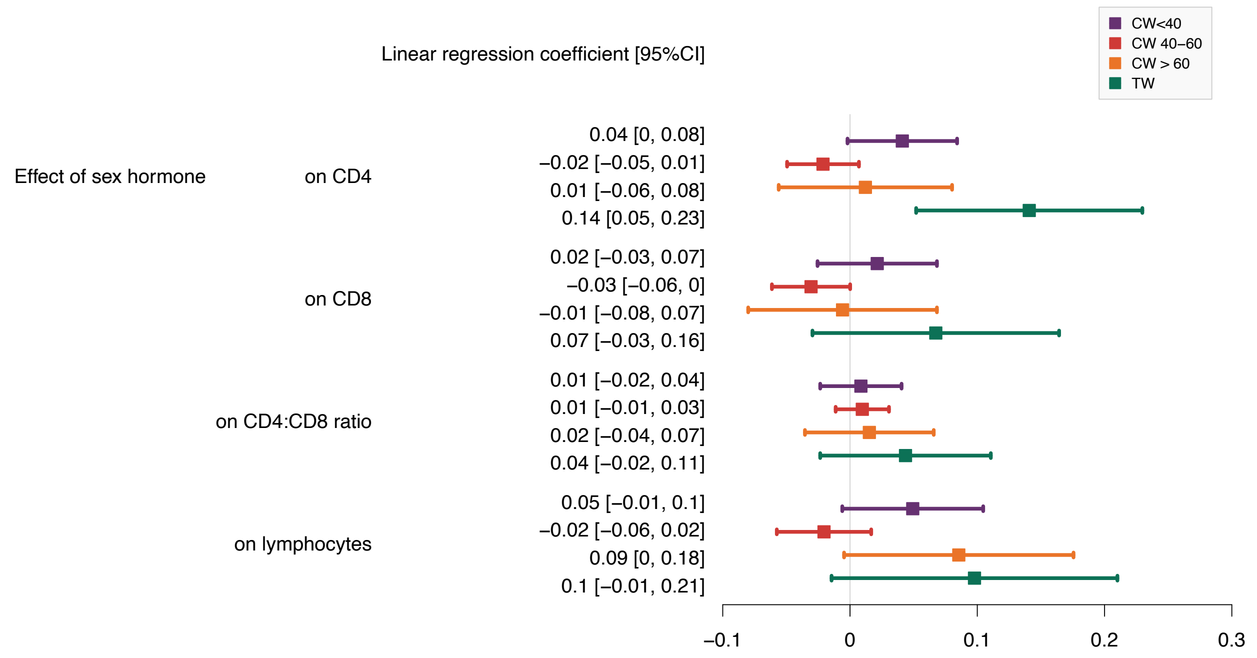

**Supplementary Figure 8.** Comparison of the effect of sex hormones in CW<40 (purple), CW 40-60 (red), CW>60 (orange), and TW (green) and the difference between CW and CM (dark grey) on the studied immune markers

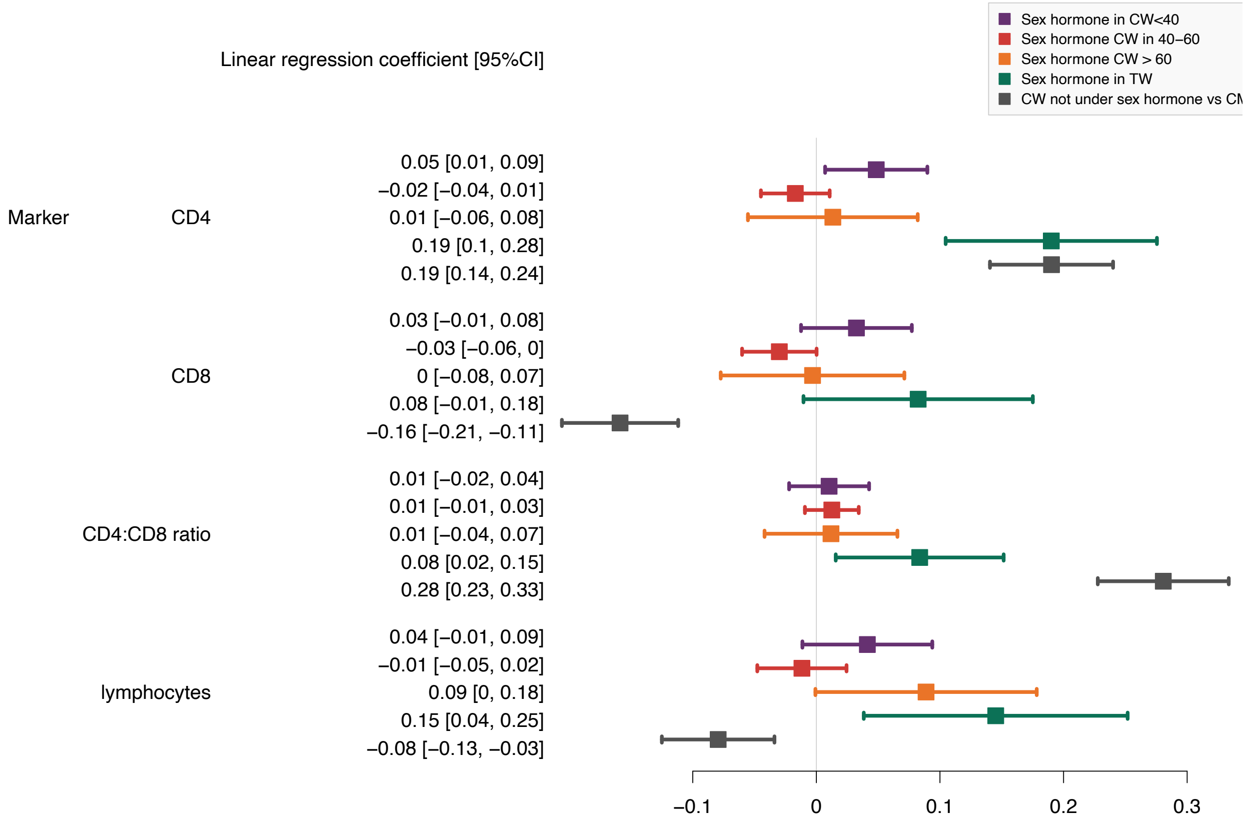

**Supplementary Figure 9.** Heatmap representing the normalized protein expression (NPX) in the 62 studied plasma samples and their classification in two groups: before sex hormone intake (grey) and after sex hormone intake (purple).

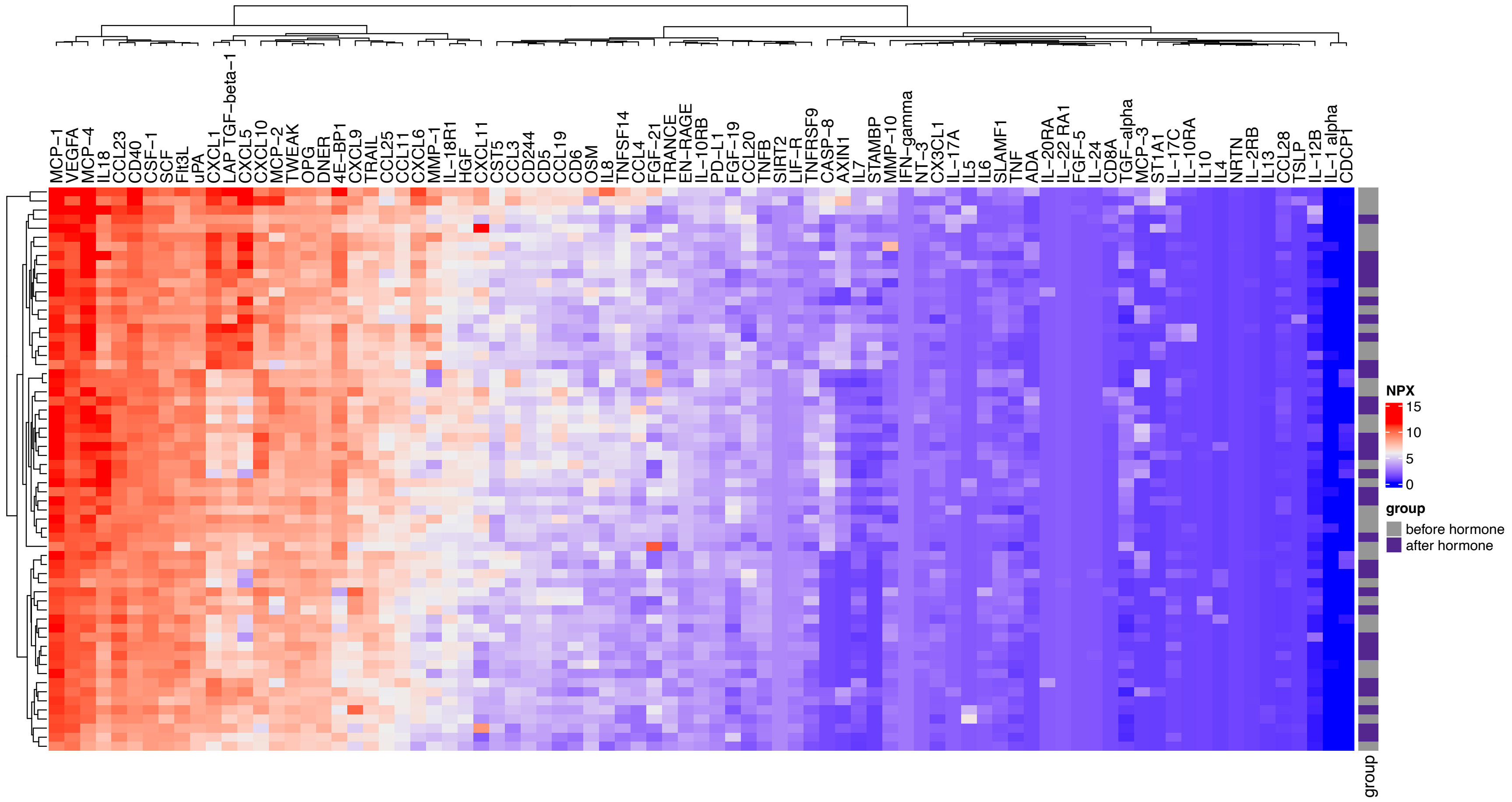

**Supplementary Figure 10.** Normalized protein expression (NPX) of NT-3 and STAMBP in plasma samples before sex hormone intake (grey) and after (purple). Boxplots represent the following: median with the middle line, upper and lower quartiles with the box limits, 1.5x interquartile ranges with the whiskers.

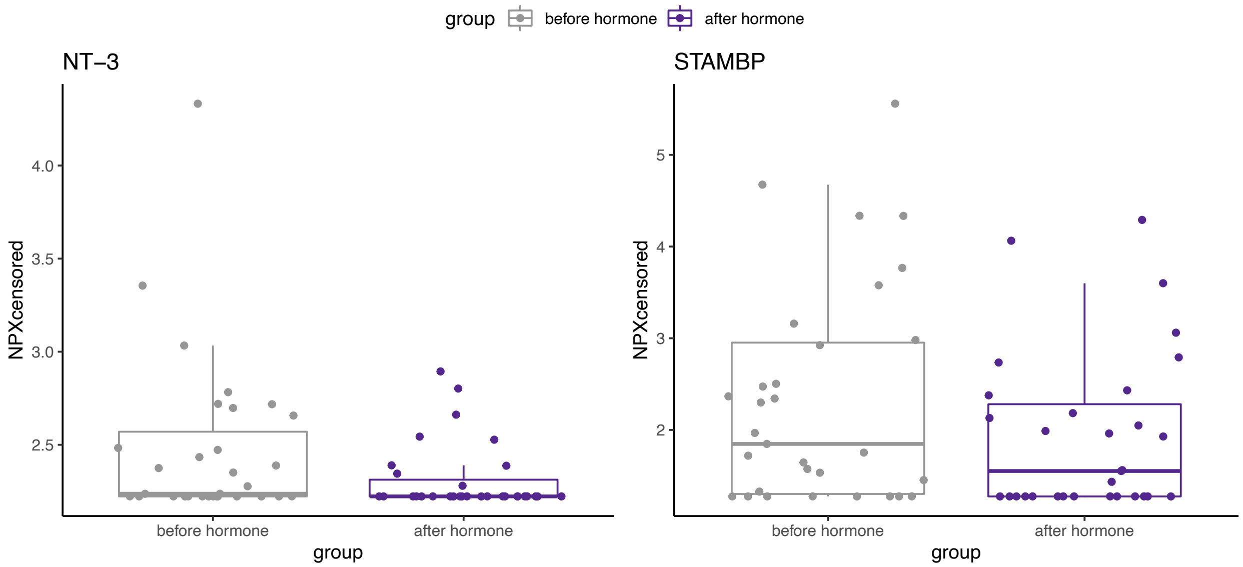

**Supplementary table 1:** Self-reported adherence, viral load (copies/mL), and depression in the different studied groups, with sex hormone intake or not.

| | CW < 40 | | CW 40-60 | | CW > 60yo | | TW | |
|---|---|---|---|---|---|---|---|---|
| **Under sex hormones** | No | Yes | No | Yes | No | Yes | No | Yes |
| | (N=7968) | (N=1115) | (N=26666) | (N=2781) | (N=4981) | (N=546) | (N=521) | (N=669) |
| **Self-reported adherence** | | | | | | | | |
| never missed | 6336 (79.5%) | 930 (83.4%) | 22626 (84.8%) | 2424 (87.2%) | 4533 (91.0%) | 504 (92.3%) | 456 (87.5%) | 600 (89.7%) |
| missed once a month | 1008 (12.7%) | 131 (11.7%) | 2633 (9.9%) | 284 (10.2%) | 302 (6.1%) | 28 (5.1%) | 50 (9.6%) | 48 (7.2%) |
| missed more than once every 2 weeks | 624 (7.8%) | 54 (4.8%) | 1407 (5.3%) | 73 (2.6%) | 146 (2.9%) | 14 (2.6%) | 15 (2.9%) | 21 (3.1%) |
| **VL** | | | | | | | | |
| <50 | 7445 (93.4%) | 1075 (96.4%) | 25512 (95.7%) | 2697 (97.0%) | 4773 (95.8%) | 545 (99.8%) | 475 (91.2%) | 659 (98.5%) |
| 50-1000 | 379 (4.8%) | 32 (2.9%) | 904 (3.4%) | 60 (2.2%) | 182 (3.7%) | 0 (0%) | 39 (7.5%) | 9 (1.3%) |
| >1000 | 144 (1.8%) | 8 (0.7%) | 250 (0.9%) | 24 (0.9%) | 26 (0.5%) | 1 (0.2%) | 7 (1.3%) | 1 (0.1%) |
| **Depression** | | | | | | | | |
| No | 7082 (88.9%) | 965 (86.5%) | 22334 (83.8%) | 2326 (83.6%) | 4207 (84.5%) | 444 (81.3%) | 402 (77.2%) | 503 (75.2%) |
| Yes | 886 (11.1%) | 150 (13.5%) | 4332 (16.2%) | 455 (16.4%) | 774 (15.5%) | 102 (18.7%) | 119 (22.8%) | 166 (24.8%) |

**Supplementary Table 2.** Comparison between protein concentration before and after sex hormone levels. For each protein, we display the test used (paired t.test if no left-censored data, cen_paired if left-censored data), its corresponding p-value, and the adjusted p-value using FDR method.

| Assay | pval.test | pval.adjust | test |
|---|---|---|---|
| NT-3 | 0.019 | 0.760 | cen_paired |
| STAMBP | 0.032 | 0.760 | cen_paired |
| CD244 | 0.064 | 0.760 | t.test |
| CD6 | 0.066 | 0.760 | t.test |
| IL6 | 0.070 | 0.760 | cen_paired |
| IL7 | 0.084 | 0.760 | cen_paired |
| CASP-8 | 0.091 | 0.760 | cen_paired |
| MCP-4 | 0.100 | 0.760 | t.test |
| TNFSF14 | 0.109 | 0.760 | t.test |
| IL18 | 0.113 | 0.760 | t.test |
| AXIN1 | 0.142 | 0.841 | cen_paired |
| CCL19 | 0.165 | 0.841 | t.test |
| IL-17C | 0.193 | 0.841 | cen_paired |
| CCL4 | 0.207 | 0.841 | cen_paired |
| LAP TGF-beta-1 | 0.208 | 0.841 | t.test |
| TGF-alpha | 0.264 | 0.841 | t.test |
| MCP-1 | 0.265 | 0.841 | t.test |
| CXCL11 | 0.277 | 0.841 | t.test |
| CXCL6 | 0.291 | 0.841 | t.test |
| uPA | 0.312 | 0.841 | t.test |
| OSM | 0.316 | 0.841 | t.test |
| MMP-10 | 0.350 | 0.841 | t.test |
| CD40 | 0.352 | 0.841 | t.test |
| TNFRSF9 | 0.366 | 0.841 | t.test |
| CD5 | 0.368 | 0.841 | t.test |
| ST1A1 | 0.383 | 0.841 | cen_paired |
| CXCL5 | 0.396 | 0.841 | t.test |
| IL-10RA | 0.398 | 0.841 | cen_paired |
| CXCL1 | 0.420 | 0.841 | t.test |
| 4E-BP1 | 0.429 | 0.841 | t.test |
| CCL20 | 0.430 | 0.841 | t.test |
| OPG | 0.439 | 0.841 | t.test |
| IL8 | 0.440 | 0.841 | t.test |
| FGF-21 | 0.456 | 0.841 | cen_paired |
| HGF | 0.466 | 0.841 | t.test |
| CCL11 | 0.467 | 0.841 | t.test |
| ADA | 0.468 | 0.841 | cen_paired |
| CX3CL1 | 0.483 | 0.841 | t.test |
| CSF-1 | 0.497 | 0.841 | t.test |
| TNF | 0.502 | 0.841 | t.test |
| TRAIL | 0.528 | 0.850 | t.test |
| CCL3 | 0.549 | 0.850 | t.test |
| MCP-2 | 0.555 | 0.850 | t.test |
| PD-L1 | 0.558 | 0.850 | t.test |
| DNER | 0.581 | 0.866 | t.test |
| TNFB | 0.602 | 0.866 | t.test |
| EN-RAGE | 0.608 | 0.866 | t.test |
| VEGFA | 0.624 | 0.871 | t.test |
| MCP-3 | 0.665 | 0.893 | cen_paired |
| CCL25 | 0.668 | 0.893 | t.test |
| CST5 | 0.688 | 0.893 | t.test |
| IL-18R1 | 0.702 | 0.893 | t.test |
| LIF-R | 0.723 | 0.893 | t.test |
| SCF | 0.739 | 0.893 | t.test |
| Flt3L | 0.741 | 0.893 | t.test |
| IL-17A | 0.759 | 0.893 | cen_paired |
| CCL23 | 0.774 | 0.893 | t.test |
| TWEAK | 0.781 | 0.893 | t.test |
| IL-12B | 0.792 | 0.893 | cen_paired |
| MMP-1 | 0.807 | 0.893 | t.test |
| CCL28 | 0.813 | 0.893 | cen_paired |
| SLAMF1 | 0.862 | 0.931 | t.test |
| IL-10RB | 0.886 | 0.942 | t.test |
| CXCL10 | 0.906 | 0.943 | t.test |
| CXCL9 | 0.915 | 0.943 | t.test |
| TRANCE | 0.958 | 0.965 | t.test |
| FGF-19 | 0.965 | 0.965 | cen_paired |
